# Prevalence of Fetal Alcohol Spectrum Disorder (FASD) in Greater Manchester, UK: an active case ascertainment study

**DOI:** 10.1101/2021.05.28.21257704

**Authors:** Robyn McCarthy, Raja A.S. Mukherjee, Kate M. Fleming, Jonathan Green, Jill Clayton-Smith, Alan D. Price, Clare S. Allely, Penny A. Cook

## Abstract

**Background:** Despite high levels of prenatal alcohol exposure in the UK, evidence on the prevalence of fetal alcohol spectrum disorders (FASD) is lacking. This paper reports on FASD prevalence in a small sample of children in primary school.

**Methods:** A two-phase active case ascertainment study was conducted in three mainstream primary schools in Greater Manchester, UK. Schools were located in areas that ranged from relatively deprived to relatively affluent. Initial screening of children aged 8-9 years used pre-specified criteria for elevated FASD risk (small for age; special educational needs; currently/previously in care; significant social/emotional/mental health symptoms). Screen positive children were invited for detailed ascertainment of FASD using gold standard measures including medical history, facial dysmorphology, neurological impairment, executive function and behavioural difficulties.

**Results:** Of 220 eligible children, 50 (23%) screened positive and 12% (26/220) proceeded to phase-two assessment. Twenty had a developmental disorder, of which, four had FASD and four were assessed as possible FASD. The crude prevalence rate of FASD in these schools was 1.8% (95%CI: 1.0%,3.4%) and when including possible cases was 3.6% (2.1%,6.3%). None of these children had previously identified with a developmental diagnosis.

**Conclusions:** FASD was found to be common in these schools, but limitations to the sampling restrict inferences to a population prevalence. Most of these children’s needs had not previously been identified.

## Introduction

An estimated 10% of pregnancies globally are exposed to alcohol, a potent teratogen that can lead to physical and neurodevelopmental birth defects (Popova et al., 2017), collectively known as Fetal Alcohol Spectrum Disorders (FASD) (Cook et al., 2016). FASD is an umbrella term that includes the diagnoses of Fetal Alcohol Syndrome (FAS), pFAS (partial Fetal Alcohol Syndrome), ARBD (Alcohol-Related Birth Defects), and ARND (Alcohol-Related Neurodevelopmental Disorder)/ND-PAE (Neurobehavioral Disorder Associated with Prenatal Alcohol Exposure).

The prevalence of FASD is estimated to be 0.8% globally and highest in Europe, at 2% (Lange et al., 2017a). There are no direct estimates of prevalence in the four countries with the highest known rates of prenatal alcohol exposure (Ireland, Belarus, Denmark and UK), all of which have rates of over 40% pregnancies exposed to alcohol (Popova et al., 2017). For the UK, the modelled estimate suggests 3.2% of children and young people may have FASD (Lange et al., 2017a). A national study in the US, on populations relatively similar to those in the UK, found a weighted estimate of 3-10% for FASD in children in primary school (May et al., 2018). Where a UK cohort with high levels of exposure (79% of mothers drank during the pregnancy, with 25% at binge levels) was used, 6–17% of children screened positive for features of FASD (McQuire et al., 2019). However, the lack of direct evidence of prevalence contributes to under-investment in diagnostic, treatment and prevention services (Scholin et al., 2021).

Active case ascertainment studies, the basis of global and national prevalence estimates (Lange et al., 2017a; May et al., 2018), are considered the ‘gold standard’ method of estimating prevalence, and involve screening a cross section of the general population of children (Roozen et al., 2016). Passive methods are less useful because individuals with FASD are often not diagnosed (May & Gossage, 2001; Morleo et al., 2011) for a number of reasons, including a lack of knowledge/training among healthcare/educational professionals (Mukherjee et al., 2015); difficulty in differentiating features of FASD from other commonly co-occurring disorders (e.g. Attention-Deficit/Hyperactivity Disorder, ADHD, or Autism Spectrum Disorder, ASD) (Chasnoff, Wells & King, 2015; Mukherjee et al., 2011; Young et al., 2016).

The aim of this study was to provide direct evidence of the prevalence of FASD in a small sample of UK children aged 8 to 9 years.

## Materials and Methods

### Setting

The setting was Greater Manchester, North West England (population 2.8 million), an area with a higher than England average level of alcohol harm (Public Health England, 2021), and relatively high deprivation (Ministry of Housing Communities and Local Government, 2019).

This study was part of a wider initiative, the ‘Preventing Alcohol Exposed Pregnancy Programme’ taking place in four of the ten Greater Manchester local authority areas. The initiative also included increased awareness raising and interventions with women who were pregnant or at risk of unplanned pregnancy (Reynolds et al., 2021).

### Design

We report on a cross-sectional study to detect cognitive impairments and associated conditions, including FASD, using an Active Case Ascertainment method. Based on the World Health Organization (WHO) standard protocol for FASD prevalence studies (World Health Organisation, 2012), children in school year three/four (8-9 years of age at enrolment) who were able to communicate in English were invited to take part in a two-phase approach. Children with a known risk factor for FASD went to the second phase ‘Children with a known risk factor for FASD went to the second phase (full assessment, see section below, phase 1: initial screening, Figure 1). Child assessments took place between July 2019 and March 2020. Restrictions related to the COVID-19 pandemic prevented face to face data collection from mid-March 2020, at which point there was one outstanding parent interview, which was conducted by telephone in March 2020.

**Figure 1.**
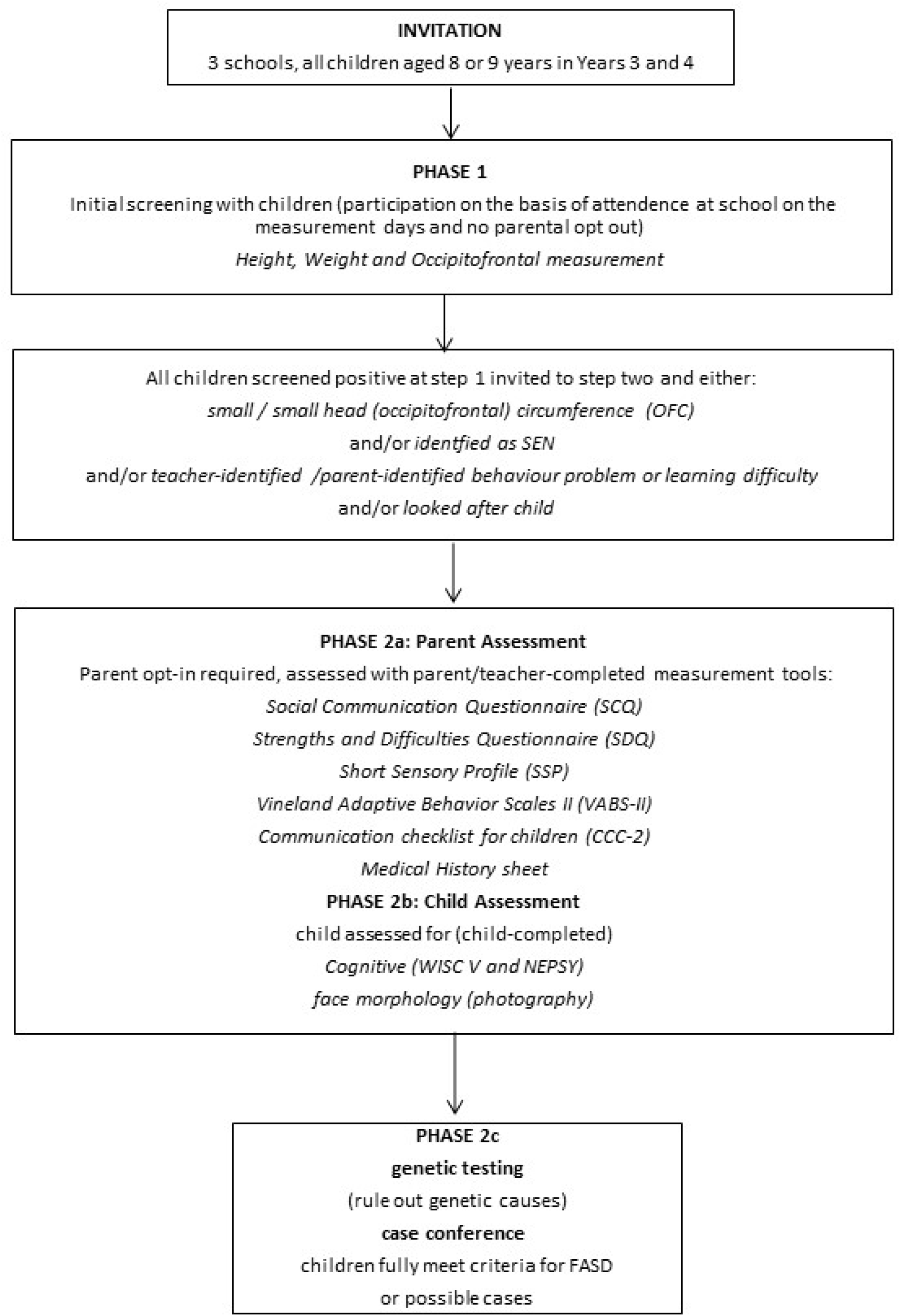
Study design

### Sample size

A sample size of 170 was adequate to get a preliminary indication of prevalence in a selected sample of schools (based on estimated true prevalence=0.03, precision=0.05; sensitivity=0.85; specificity=0.85). We aimed to recruit three schools (assuming approximately 60 pupils in the relevant age category per school).

### School recruitment

In this pragmatic, small-scale study, schools were purposefully selected. Key informants (e.g. local government officials who work with schools, Education Psychologists) were consulted and suggested four schools, all of which agreed to take part. One school was excluded due to disengagement from the study and deviation from the protocol (see supplementary file 1 for details of a partial dataset). We initially aimed to include a specialist school providing Social Emotional and Mental Health (SEMH) support. All such schools within the study area were contacted. One such school agreed to take part but withdrew after being unable to gain consent from any parents.

In February 2020, an extension of data collection was granted with the aim to recruit further schools to increase the baseline sample size. Four additional schools were recruited. However, the imposition of COVID-19 related restrictions, including lockdown, meant that the study could not be completed in the second wave of schools. Partial results are reported in Supplementary file 1.

### Phase 1: Initial Screening

Parents of all eligible children were sent a letter describing the study, with the option to opt out, at least five days before data collection. Parents who wished to remove their child from the study at this stage were advised to return the letter with the opt-out option selected. Researchers were present in school to assess physical measurements. Height was measured using a mobile stadiometer, weight using Marsden MBF-6000 scales and head circumference (OFC) using a Sec 201 measuring tape. To become eligible for phase 2 (full assessment), one or more of the following criteria had to be met: height or weight below the ninth centile or OFC below the second centile; identified by the school or parents as having difficulties with learning, maladaptive behaviour, inattention or hyperactivity issues; have an Education Health and Care plan; be a currently or previously looked after child; already have diagnosed difficulties with behaviour including ADHD or conduct disorder. Exclusion criteria: disabilities or behavioural abnormalities known to be caused by well-characterised and already identified genetic factors (e.g. Down Syndrome, Williams Syndrome) or by post-natal brain injuries. These were excluded because the physical/behavioural stigmata overlap with FASD in presentation, and an FASD label cannot be attributed as the primary aetiological factor.

### Phase 2a: Measures completed by parents/carers

Parents of those meeting criteria for phase 2 were sent information sheets and consent forms to opt themselves and their child into the study either by email, letter sent home, by telephone or in person by the Special Educational Needs Coordinator or head teacher. Where children were currently under the care of the local authority consent was obtained from the supervising social worker. For parent-report assessments, most took place at school in a private room for two-hour sessions. Where requested by the parent, assessments took place in their home.

Parent report assessments used the validated measures listed in Figure 1. The medical history was taken using a structured questionnaire developed originally at the University of New Mexico (May et al., 2018). The schedule included questions on pre-pregnancy, pregnancy and current substance use (alcohol, tobacco, prescription and illicit drugs), folic acid use in pregnancy and birth complications. General questions on current alcohol consumption were asked first as part of wider lifestyle set of questions, followed by questions about alcohol consumption in pregnancy. If alcohol was consumed during pregnancy, further questions ascertained the level and timing of consumption. Birth mothers were interviewed privately in order to encourage open reporting of alcohol use. For looked after children, fostered or adopted children, the parent/carer with most knowledge of the child was interviewed.

### Phase 2b: Measurements on children

Measurements took place in school during school hours. Dysmorphology of fetal alcohol syndrome (FAS) facial characteristics was assessed from photographs (using standardised alignment of the participant’s head relative to the camera lens) taken by trained researchers. Images were analysed by FAS Facial Photographic Analysis Software (Astley, 2015) and validated by an experienced clinical geneticist. Neurological impairment was assessed using the Developmental NEuroPSYchological Assessment (NEPSY) to assess memory, attention, and executive functions. The NEPSY subtests (Inhibition, Narrative Memory and Word List Interference, Animal Sorting, Clocks and Memory for Faces) were those that had been identified in previous FASD research (Rasmussen et al., 2013). The Wechsler Intelligence Scale for Children (WISC V) (Kaufman et al., 2015) was used to assess the child’s cognition, as used in previous FASD studies (Raldiris et al., 2018).

### Phase 2c: Ruling out genetic causes and case conference

The results of the assessments were compared to the case definition (Table 1), derived from the WHO protocol for prevalence studies (World Health Organisation, 2012) and international guidelines (Cook et al., 2016). Where deficits met criteria in three subdomains, and PAE was present, FASD was considered. Where a child had all three sentinel facial features associated with FAS, FASD was considered in the absence of reported PAE. For all those considered for FASD, a Microarray Comparative Genomic Hybridisation (array CGH) test was used to rule out other disorders that may present with a similar behavioural presentation of genetic origin (Douzgou et al., 2012). A saliva sample was taken using the Oragene DNA OG-575 (DNAGenotek, 2020). The Oligo (Oxford Gene Technology, Oxford, UK) Hx60k array was carried out by the North Western Regional Genetics Laboratory within Manchester University NHS Foundation Trust, Manchester.

**Table 1:**
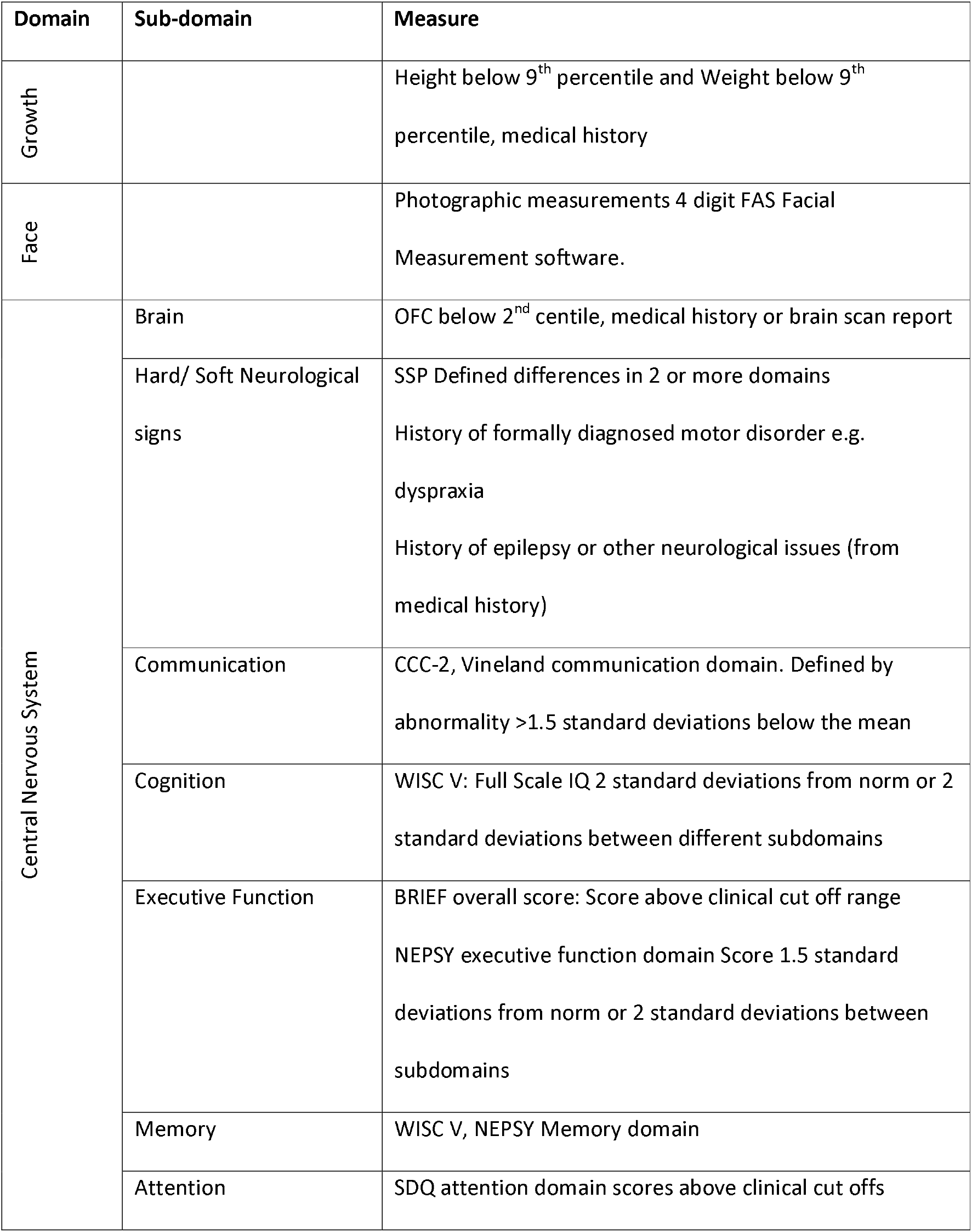
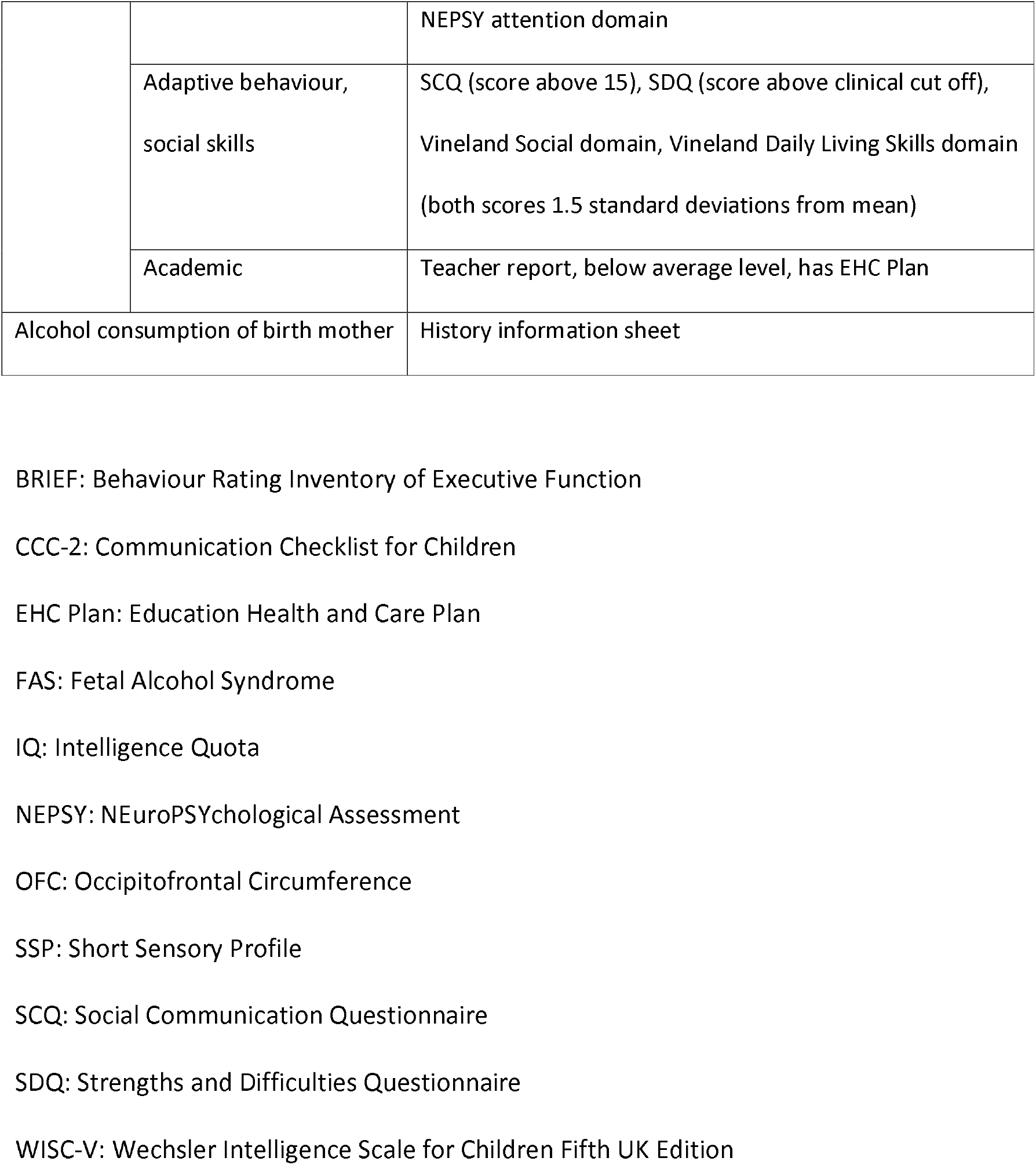
Measures used and Fetal Alcohol Spectrum Disorder (FASD) domains of impairment.

| Domain | Sub-domain | Measure |
| --- | --- | --- |
| Growth |  | Height below 9<sup>th</sup> percentile and Weight below 9<sup>th</sup> percentile, medical history |
| Face |  | Photographic measurements 4 digit FAS Facial Measurement software. |
| Central Nervous System | Brain | OFC below 2<sup>nd</sup> centile, medical history or brain scan report |
|  | Hard/ Soft Neurological signs | SSP Defined differences in 2 or more domains<br>History of formally diagnosed motor disorder e.g. dyspraxia<br>History of epilepsy or other neurological issues (from medical history) |
|  | Communication | CCC-2, Vineland communication domain. Defined by abnormality >1.5 standard deviations below the mean |
|  | Cognition | WISC V: Full Scale IQ 2 standard deviations from norm or 2 standard deviations between different subdomains |
|  | Executive Function | BRIEF overall score: Score above clinical cut off range<br>NEPSY executive function domain Score 1.5 standard deviations from norm or 2 standard deviations between subdomains |
|  | Memory | WISC V, NEPSY Memory domain |
|  | Attention | SDQ attention domain scores above clinical cut offs |
|  |  | NEPSY attention domain |
|  | Adaptive behaviour,<br>social skills | SCQ (score above 15), SDQ (score above clinical cut off),<br>Vineland Social domain, Vineland Daily Living Skills domain<br>(both scores 1.5 standard deviations from mean) |
|  | Academic | Teacher report, below average level, has EHC Plan |
| Alcohol consumption of birth mother |  | History information sheet |

BRIEF: Behaviour Rating Inventory of Executive Function
CCC-2: Communication Checklist for Children
EHC Plan: Education Health and Care Plan
FAS: Fetal Alcohol Syndrome
IQ: Intelligence Quota
NEPSY: NEuroPSYchological Assessment
OFC: Occipitofrontal Circumference
SSP: Short Sensory Profile
SCQ: Social Communication Questionnaire
SDQ: Strengths and Difficulties Questionnaire
WISC-V: Wechsler Intelligence Scale for Children Fifth UK Edition

Findings for each child discussed during case conferences attended by the study team including clinicians with expertise in the diagnosis of FASD. Cases of FASD were defined according to internationally recognised guidelines (Cook et al., 2016). Possible cases were where FASD was suspected but information was missing or unclear, for example regarding alcohol use.

### Data analysis

Prevalence was estimated as the total number of children with FASD and probable FASD as the numerator, and the total number of eligible children at the same site as the denominator. This generates a conservative or minimum estimate of prevalence as it assumed that all children who were not examined did not have FASD. To obtain confidence intervals we accounted for clusters (schools) by using nonparametric bootstrapping, resampling with replacement clusters with 10000 bootstrap runs.

### Ethical considerations

Parents were invited to take part and were offered a report on their child’s learning and behaviour issues, which could include a range of learning difficulties, including possible ASD, ADHD and FASD. The text of the information sheet made it clear that the main objective was to ascertain FASD prevalence.

A small number of participants took part in genetic testing, which can occasionally identify markers for other health conditions. The information sheet sought to ensure that the parents/carers understood possible outcomes of the genetic test before giving consent.

Ethical approval was granted by the University of Salford Ethics committee in May 2019 (reference: HSR1819-100).

## Results

### Participating schools

Schools represented a range of deprivation when measured by the index of multiple deprivation. School 1 was located in one of England’s 20% most deprived areas, and school 3, 30%. School 2 was located among the 20% most affluent areas in England.

### Flow of participants

A total of 220 children were invited and 203 children took part in the phase 1 physical measures (height, weight and OFC) (Figure 2). Fifty children were eligible for full assessment (phase 2) because of screening positive on the physical measures, parent/teacher concerns, already acknowledged as SEN, or currently or previously being in local authority care (Table 2).

**Figure 2.**
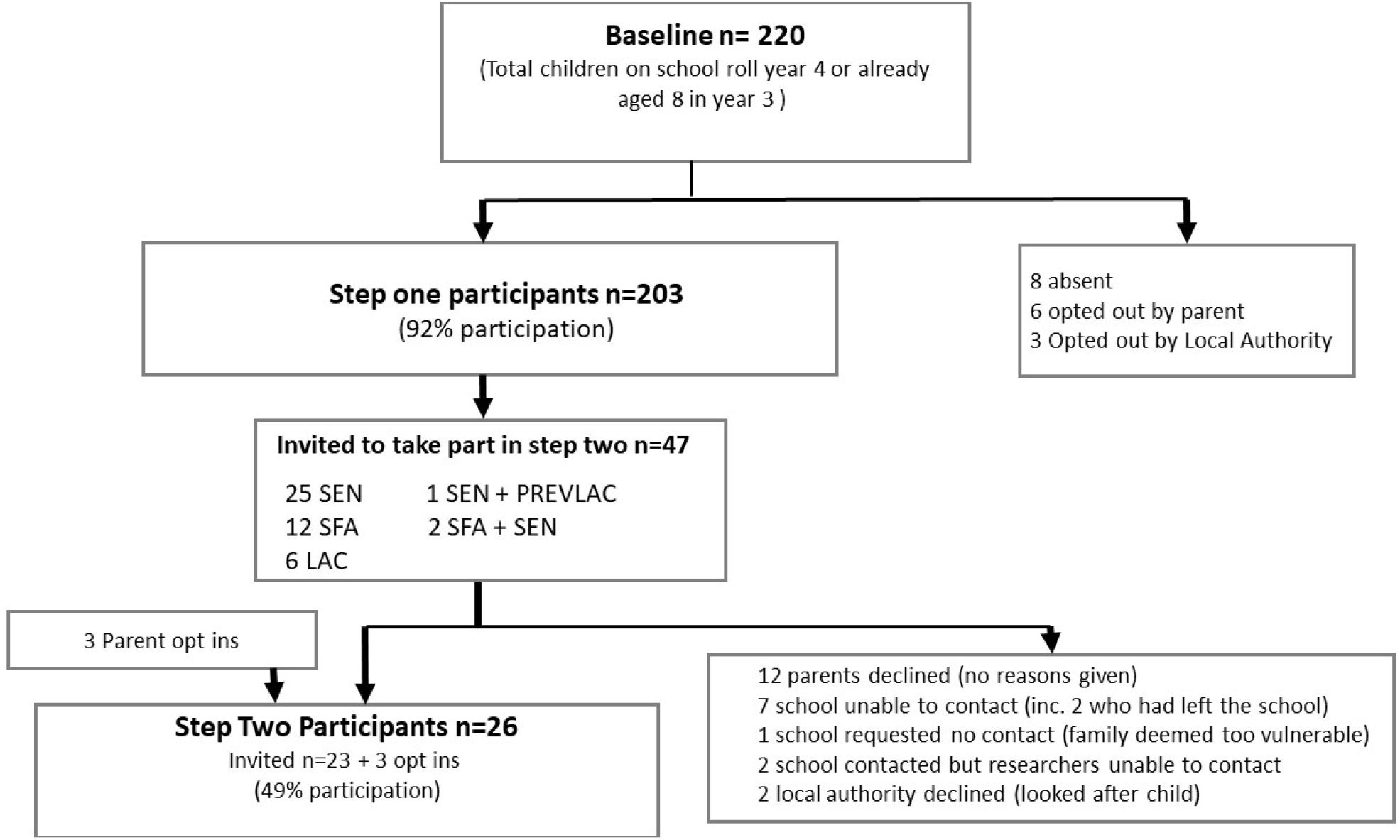
Flow of participants through the study. SFA: small for age; SEN: on school educational needs register; LAC: ‘looked after child’ i.e. in the care of the local authority; PrevLAC: previously looked after, i.e. adopted.

**Table 2:** Screening and assessment data by school.

|  | **School1**<br>**(baseline N=44)** | **School2**<br>**(baseline N=118)** | **School3**<br>**(baseline**<br>**N=59)** | **Total (N=220)** |
| --- | --- | --- | --- | --- |
| **Screened at stage one** n<br>(% of baseline) | 38 (86) | 109 (92) | 56 (95) | 203 (92) |
| **Sex**, n(% of baseline) |  |  |  |  |
| Male | 18 (40) | 55 (46) | 30 (51) | 103 (47) |
| Female | 20 (45) | 54 (46) | 26 (44) | 100 (45) |
| **Age**, Years (y) & months<br>(m) |  |  |  |  |
| Median | 8y11m | 8y10m | 8y6m | 8y10m |
| [Interquartile range] | [8y9m, 9y5m] | [8y5m, 9y6m] | [8y3m, 8y11m] | [8y 5m, 9y4m] |
| (range) | (8y3m, 9y9m) | (8y0m, 9y10m) | (8y0m, 9y8y) | (8y0m, 9y10m) |
| **Physical screen positive**<br>n (% of baseline) |  |  |  |  |
| Height <9<sup>th</sup> percentile | 4 (9) | 7 (6) | 2 (3) | 13 (6) |
| Weight <9<sup>th</sup> percentile | 2 (5) | 3 (3) | 2 (3) | 7 (3) |
| OFC <2th percentile | 1 (2) | 1(1) | 1 (2) | 3 (1) |
| **Reason for recruitment** n (% of invited) |  |  |  |  |
| SEN | 14(66) | 7 (43) | 5 (50) | 26 (13) |
| SFA | 3(14) | 5 (31) | 4 (40) | 12 (6) |
| LAC | 2(10) | 3 (18) | 1 (10) | 6 (3) |
| SFA+SEN | 2(10) | 0 | 0 | 2 (1) |
| SFA + prevLAC | 0 | 1 (6) | 0 | 1 (0.5) |
| Parent opt-in | 0 | 3 (16) | 0 | 3 (1) |
| **Invited to second phase** | 21 | 19 | 10 | 50 |
| **Phase two assessments** | **N=10** | **N=9** | **N=7** | **N=26** |
| Referral reason (% of phase<br>two participants) |  |  |  |  |
| SEN | 5 (50) | 3 (33) | 4 (57) | 12 (46) |
| SFA | 3 (30) | 0 | 2 (29) | 5 (19) |
| LAC | 0 | 2 (22) | 1 (14) | 3 (12) |
| SFA and SEN | 2 (20) | 0 | 0 | 2 (8) |
| SFA and PrevLAC | 0 | 1 (11) | 0 | 1 (4) |
| parent opt in | 0 | 3 (33) | 0 | 3 (11) |
| **Maternal Ethnicity**, n(%) |  |  |  |  |
| White UK | 10 (100) | 8 (89) | 6 (86) | 24 (92) |
| Non white | 0 | 1 (11) | 0 | 1 (4) |
| Not reported | 0 | 0 | 1 (14) | 1 (4) |
| **Maternal educational qualifications, highest received n(%)** |  |  |  |  |
| <4 GCSEs | 2 (20) |  | 1 (14) | 3 (12) |
| 4+GCSEs | 4 (40) |  | 2 (29) | 6 (23) |
| 2+A levels | 2 (20) | 3 (33) | 1 (14) | 6 (23) |
| Degree | 2 (20) | 4 (44) | 2 (29) | 8 (30) |
| Missing data | 0 | 2 (22) | 1 (14) | 3 (12) |
| **Full assessment** |  |  |  |  |
| **Deficits in three subdomains met, n(% of assessed)** | 8 (80) | 7 (78) | 6 (86) | 21 (80) |
| **FASD, n (% of assessed)** |  |  |  |  |
| FASD | 1 (10) | 3 (33) | 0 (0) | 4 (15) |
| FASD and possible FASD | 3 (30) | 4 (44) | 1 (10) | 8 (30) |
| **Other outcomes** |  |  |  |  |
| ASD | 0 (0) | 2 (22) | 3 (43) | 5 (19) |
| ADHD | 2 (20) | 0 (0) | 1 (14) | 3 (12) |
| DLD | 2 (20) | 1 (10) | 0 (0) | 3 (12) |
| General learning disability | 0 (0) | 0 (0) | 2 (28) | 2 (8) |
| No disorder | 3 (30) | 2 (20) | 0 (0) | 5 (19) |
| **FASD prevalence per 100 [95% CI]** |  |  |  |  |
| FASD | 2.3 [0.3,14.8] | 2.5 [0.8, 7.4] | No observations | 1.8 [1.0,3.4] |
| FASD and possible FASD | 7.0 [2.2,19.8] | 3.4 [1.3, 8.7] | 1.7 [0.2,11.1] | 3.6 [2.1,6.3] |

SEN: on school educational needs register; SFA: small for age; LAC: 'looked after child', i.e.
under care of local authority; PrevLAC: previously looked after child, i.e. adopted; OFC:
occipital frontal circumference; GCSE: General Certificate of Education, qualification usually
taken at age 16 years; A level: Advanced Level, qualification usually taken at age 18 years;
FASD: fetal alcohol spectrum disorder; ASD: autism spectrum disorder; ADHD: attention
deficit hyperactivity disorder; DLD: developmental language disorder.

Assessment was not possible on 24 children due largely to parents not giving consent or being uncontactable.

Full assessment took place with 26 children, four of whom had FASD and a further four were identified as possible FASD (Table 2). Other developmental disorders identified are detailed in supplementary file 2.

Of the children with full data sets, none had reported exposure to known teratogenic antiepileptic pharmaceuticals. Two had exposure to illegal drugs and over two thirds of the children assessed (68%) has some prenatal exposure to alcohol (Table 3).

**Table 3:** prenatal exposure data from cases with a complete dataset.

| **Reported Prenatal exposures** | **FASD**<br>**N=4** | **Possible**<br>**FASD n=4** | **Other**<br>**N= 18** | **TOTAL**<br>**N=26** |
| --- | --- | --- | --- | --- |
| Anticonvulsants | 0 | 0 | 0 | 0 |
| Illegal drugs | 1(25%) | 0 | 1 (6%) | 2 (8%) |
| Alcohol |  |  |  |  |
| Significant risk<sup>a</sup> | 3 (75%) | 3(75%) | 4 (22%) | 10 (38%) |
| Low risk<sup>b</sup> | 0 | 1 (25%) | 7 (40%) | 8 (30%) |
| No alcohol reported | 1(25%) | 0 | 7 (40%) | 8 (30%) |

<sup>a</sup>Cut off for significant risk defined as four or more alcoholic drinks on four or more
occasions or greater than seven units a week throughout the term of the pregnancy
<sup>b</sup> Low risk: alcohol consumed, but not meeting threshold above (Kelly, et al., 2009; Sayal et al., 2013).

Of the FASD cases, none had a prior clinical diagnosis of any neurodevelopmental disorder or an Education, Health and Care (EHC) Plan, and only one child had been identified by the school as having special educational needs. Three out of the four cases had high risk prenatal alcohol exposure reported, while in the fourth, alcohol was not reported but the child had severe FAS facial features, was small for age, a low full scale IQ of 66, and a normal CGH array (a summary of cases and possible cases assessed against the FASD criteria is given in supplementary file 2).

Of the possible cases where FASD was suspected but not confirmed, none had a clinical diagnosis of any neurodevelopmental disorder and only one had an EHC Plan. All were on the schools’ special educational needs register, had prenatal exposure to alcohol, and had deficits in at least three subdomains. Of the 12 children with a suspected disorder that was deemed unlikely to be FASD, 11 had some level of prenatal exposure to alcohol (Table 3).

### Prevalence Rates

We calculated a conservative (minimum) prevalence of FASD of 1.8% 4/220 (95% CI [1.0, 3.4]), and a conservative (minimum) prevalence that also included possible FASD of 3.6% 8/220 (95%CI [2.1, 6.3]) within our study population.

## Discussion

### Prevalence of FASD

This is the first FASD active case ascertainment study to be carried out in the UK, a country with one of the highest rates of drinking in pregnancy in the world. These prevalence estimates, though not necessarily generalisable to other communities, are in line with a modelled population prevalence estimate for the UK of 3.2% (Lange et al., 2017b) and consistent with a screening prevalence of 6-17% in a secondary analysis of data from a cohort study of children born in the 1990s (McQuire et al., 2019). As per May et al. (2018), our prevalence estimates could be considered as ‘conservative’ because we assumed all those who we did not fully assess did not have FASD. Further work examining the prevalence of FASD taking account of maternal sociodemographic factors and trends in alcohol consumption would allow a more precise estimate of the likely burden of this condition in the UK.

The study region (Greater Manchester) sees approximately 34,000 babies born per year (ONS, 2020), yet diagnoses only around 36 cases of FASD per year (unpublished data from a Freedom of Information request). This low rate of diagnosis in relation to the example local prevalence we have demonstrated here suggests that increased recognition and diagnostic capacity is required. This is important because early diagnosis and support for families affected by FASD can prevent or mitigate adverse secondary outcomes, such as school exclusions, poor job prospects and mental ill-health (Alex & Feldmann, 2012; Landgren, et al., 2019; Rangmar, et al., 2016Streissguth, et al., 2004).

### Strengths and Limitations

We demonstrated successful engagement with the three schools who made intensive efforts on our behalf to recruit parents. The schools covered a range of communities in different levels of deprivation, with two serving relatively deprived populations and one relatively affluent. A larger, more definitive study would use a random sampling technique, stratified by deprivation level, to select schools.

We were unable to obtain sufficient information for almost half (24/50) of the children who were identified as being at higher risk of FASD (i.e. those who were identified in phase 1 screening). A further nine were actively withdrawn before phase 1 screening and selection took place. It is not possible to know whether the proportion of cases would have been higher or lower in these groups that did not take part. It is also possible that there were cases of FASD in the remaining 161 children with no apparent risk factors. It is possible the shame and stigma associated with FASD or developmental disorders in general may have impacted on participation. Low participation has been a documented issue for other prevalence studies (Caccanti et al,. 2014; May et al., 2011; Okuliez-Kozaryn, et al., 2017) in the European region.

Extensive effort was made to contact each parent, and for those not taking part, the reasons were documented (see reasons given in Figure 2). The opportunity to have children’s needs assessed was the major incentive for schools to take part in the study. For children in the care of foster parents, consent was required from the local authority and this was particularly difficult to obtain.

We had initially hoped to include a further four schools in the prevalence calculations, however, disruptions due to COVID-19 prevented completion in these schools. Partial findings are presented in supplementary file 1.

We were not successful in collecting data in a Social Emotional and Mental Health (SEMH) school despite recruiting an enthusiastic head teacher at such a school. Anecdotally, parents felt they had nothing to gain by taking part as their child was already benefiting from specialist support. Similarly, pupil referral units were not included in this study. Together with SEMH schools, this is another setting where FASD prevalence is anticipated to be higher than the mainstream school setting.

It is likely that prenatal alcohol exposure was underestimated in this study, as dose and duration of alcohol consumption during pregnancy was calculated from retrospective self-reporting, relying on recall of behaviours taking place up to 10 years prior. Social desirability bias also affects reporting of alcohol consumption during pregnancy (Caccanti et al,. 2014, Smith et al., 2014) and we do not know how much the potential for perceived shame associated with alcohol consumption in pregnancy may have affected reporting.

It may be beneficial for future research to consider how methods could be adjusted for European populations to optimise participation and accuracy of report of alcohol consumption in these populations.

Other general limitation in studies that measure the prevalence of FASD, it that whilst there are a wide range of physical and neurological features associated with FASD, which when taken together increase the likelihood of FASD, the only features that are widely accepted as discriminating in the absence of confirmed alcohol exposure are the distinctive facial features. However, significant facial features are thought to only occur in a minority of cases of FASD, and, likewise, in this study only two out of the eight cases and possible cases showed these distinctive features. Previous prevalence studies have used a higher threshold for the physical measurements at phase 1 (e.g. a threshold of 10^th^ percentile for OFC: May et al., 2011), which would have made the screening stage more sensitive and may have led to more cases being identified. Instead, we used the Canadian diagnostic approach, where the detailed evaluation stage was more focused towards the neurocognitive deficits. We ruled out obvious genetic causes through chromosome microarray analysis and clinical geneticist review. Whilst this helped to improve confidence about the aetiological basis of the presentation, it is not possible to prove that the alcohol exposure caused the deficits. The converse also true: it is not possible to rule out alcohol as an important causal factor. Every effort was taken to reduce bias that may give rise to false positives. The case definition used validated developmental assessments and was described in advance in the study protocol and followed internationally recognised guidance (Cook et al., 2016).

An improvement on our methodology would have been to include a random sample of children for full assessment. This would have enabled us to obtain characteristics of the birth mothers in a group of children without any risk factors for FASD (i.e. those who did not meet the phase 1 screening criteria), which would have allowed more sophisticated modelling (and extrapolation) of the likely prevalence in the entire population of 8-9 year olds (May et al., 2018). In this small-scale study, we determined that this would have been difficult. Parents were prepared to take part if they thought it might benefit their child, for example by getting more information to inform their special educational needs or to obtain a diagnosis. The fact that our assessment also identified possible ASD, ADHD and other neurodevelopmental conditions was a significant ‘pull factor’ for schools and parents. It was also notable that the only children actively opted into the study because of parent concerns (and no other risk factor) were from the school in the most affluent setting (school 2). Thus, motivations to take part may differ depending on socioeconomic status. We would conclude that further research would be needed into how to incentivise parents of children with apparently typical development to take part.

### Conclusions

This is the first study in the UK to directly assess FASD in a systematically ascertained sample of children. It found FASD in 1.8% (1.0%-3.4%) of the population studied, or 3.6% (2.1%,6.3%) when possible cases were also included. Due to the small sampling frame of schools included and limitations of baseline information obtained on contacted families, we can only conclude this represents local prevalence data in typical mainstream schools rather than being able confidently to infer a ‘population prevalence’ of FASD. There are uncertainties too about this prevalence found, since half the children screened positive were lost to full ascertainment, and case identification may have been higher if all cases had been seen. Further research is needed to identify how to improve participation and accuracy of PAE in European populations.

It was already suspected from modelling and screening studies that FASD is highly prevalent in the UK (Lange et al., 2017a; McQuire et al., 2019). Confirmation that this is the case in a sample of schools should be used to increase the awareness of FASD, and invest in diagnostic services, treatment and support for those affected by FASD.

## Data Availability

Data are available from the authors upon reasonable request

## Acknowledgments

We thank the parents, guardians, children, head teachers and staff at participant schools. We also thank Róisín Reynolds and Rachael Nielsen from the Greater Manchester Health and Social Care Partnership for their support. Philip May allowed us to use the Maternal Risk Questionnaire, the University of New Mexico by the Fetal Alcohol Syndrome Epidemiology Research Team initially developed in collaboration with Sharon Wilsnack (University of North Dakota), and adapted and updated multiple times for use in prevalence studies in the United States, South Africa, and Italy. We thank those who represented families affected by FASD, led by Anna Webster. We thank the members of an independent steering group, including Anna Webster Jen Michaels, Joanne Higham, Mary Scanlon and Susan McGrail.

## Abbreviations

ADHD: Attention-Deficit/Hyperactivity Disorder
ARBD: Alcohol-Related Birth Defects
ARND: Alcohol-Related Neurodevelopmental Disorder)/
ASD: Autism Spectrum Disorder
CCC: Children’s communication Checklist
FAS: Fetal Alcohol Syndrome
FASD: Fetal Alcohol Spectrum disorders
FSIQ: Full Scale Intelligence Quotient
IDACI: Income Deprivation Affecting Children Index
IMD: Index of Multiple Deprivation
ND-PAE: Neurobehavioral Disorder Associated with Prenatal Alcohol Exposure
NEPSY: II NEuroPSYchological Assessment 2^nd^ Edition
NHS: National Health Service
OFC: Occipital Frontal Circumference
PAE: Prenatal Alcohol Exposure
pFAS: partial Fetal Alcohol Syndrome
SCQ: Social Communication Questionnaire
SDQ: Strengths and Difficulties Questionnaire
SEMH: Social, Emotional And Mental Health School
SEN: Special Educational Needs
SENCO: Special Educational Needs Coordinator
SFA: Small for age
SSP: Short sensory profile
WISC V: Weschler Intelligence scale for Children 5^th^ Edition

