## Supplementary file 1 for "Prevalence of Fetal Alcohol Spectrum Disorder (FASD) in Greater Manchester, UK: an active case ascertainment study"

**Partial dataset from schools not included in the main study**

**Recruitment and Data collection**

School Four: School four was approached by a local authority public health department. Data collection: 21 November 2019 -11 March 2020. Unsuitable rooms were provided and there was no evidence that all eligible parents had been contacted.

Schools 5-7, recruitment was via direct contact by researchers via email sent to schools in the target areas within Greater Manchester (schools 5, 6) (convenience sample); direct approach from Head Teacher (School 7, word of mouth from school 5). Data collection in schools 5 to 7 for phase 1 took place January 2020 to March 2020, before the first lockdown. Data collection in schools restarted in school 6 (15 October 2020) and School 5 and 7 (November 2020—December 2020). Modified methods were used in an attempt to continue the study during the disruption caused by COVID-19, and the study was closed down by the January 2021 lockdown. All parent interviews were conducted remotely. Child assessments were conducted in rooms with windows open and sat beside the child rather than opposite. All equipment was cleaned between sessions.

**Results**

Two children had FASD with a further two identified as possible FASD. Supplementary Table 3 shows the descriptive data for these cases. The only child opted in by a parent was found to be a possible case of FASD. Of the two children identified as FASD, neither had a prior clinical diagnosis of any neurodevelopmental disorder; and only one had an Education and Health Care plan (EHCP). Supplementary Table 4 gives the descriptive data for cases and possible cases.

**Reasons for exclusion from main study**

School 4 was excluded due to deviation from protocol. There was no evidence that all eligible children had been invited to participate in the study.

Schools 5-7 were excluded because the data collection process used modified methods and was stopped prematurely.

**Supplementary Table 1.1:** Partial dataset from screening and assessment of children in schools excluded from the main study by site

|  | **School 4** | **School 5** | **School6** | **School 7** |
| --- | --- | --- | --- | --- |
| IMD decile of school | 6 | 2 | 1 | 2 |
| Baseline N | 34 | 32 | 86 | 129 |
| Withdrawn or absent | 2 | 7 | 3 | 17 |
| **Sex**, n(% of baseline)  Male  Female | 14 (41)  17(59) | 16 (50)  16 (50) | 47 (55)  39 (45) | 69 (53)  60 (47) |
| **Age**,  years+mo (median [IQR]) | 8y5m  [8y1m-8y8m] | 8y8m  [8y2m- 8y10m] | 8y7m  [8y3m-9y2m] | 8y11m  [8y7m-9y4m] |
| **Physical screen positive** n (%)  Height <9^th^ percentile  Weight <9^th^ percentile  OFC <2^nd^ percentile  At least 1 physical screen +ve | 0  0  0  0 | 3 (9)  0  1 (3)  4 (12) | 4 (4)  3 (3)  5 (6)  8 (9) | 9 (7)  4 (3)  4 (3)  12(9) |
| **Reason for invitation** n (% of baseline) | | | | |
| SEN | 10 (29) | 4 (12) | 20 (23) | 16 (12) |
| SFA | 0 | 4 (9) | 8 (9) | 6 (5) |
| LAC  LAC+SFA | 0  0 | 0  0 | 1 (1)  0 | 1 (<1)  1 (<1) |
| SFA+SEN  Prev LAC  Parent opt-in | 0 | 1 (3) | 2 (2) | 5 (4) |
|  | 0  0 | 0  0 | 0  1 (1) | 1 (<1)  0 |
| **Invited to full assessment** | 10 (29) | 8 (25) | 29 (34) | 30 (23) |
| **Full assessment** | | | | |
| n (% of invited) | 2 (20) | 4 (50) | 8 (28) | 10 (33) |
| **Referral reason** n (% of assessed)  SEN  SFA  LAC  SFA and SEN  SFA and PrevLAC  Parent opt in | 2 (100)  0  0  0  0  0 | 2 (50)  2 (50)  0  0  0  0 | 6 (75)  1 (12)  0  0  0  1 (12) | 5 (50)  0  2 (20)  3 (30)  0  0 |
| **Maternal Ethnicity**, n (% of assessed)  White UK  Non white  Not reported | 1 (50)  1 (50)  0 | 2 (50)  2 (50)  0 | 7 (87)  1 (12)  0 | 8 (80)  2 (20)  0 |
| **Maternal educational qualifications, highest received** n (% of assessed)  <4 GCSEs  4+GCSEs  2+A levels  Degree  Missing data | 0  0  1 (50)  1 (50)  0 | 1 (25)  0  3 (75)  0  0 | 1 (12)  1 (12)  4 (50)  1 (12)  0 | 3 (30)  1 (10)  3 (30)  1 (10)  2 (20) |
| **Deficits in three subdomains met,** n (% of assessed) | 2 (100) | 2 (50) | 7 (87) | 8 (80) |
| **FASD**, n (% of assessed)  FASD  FASD and Possible FASD | 0  0 | 0  0 | 1 (12)  3 (38) | 1 (10)  1 (10) |
| **Other outcomes**  Complex (ASD/ADHD/LD)  ASD  ADHD  DLD  General learning difficulty  Nonspecific  No disorder | 2 (100) | 2 (50)  2 (50) | 2 (25)  2 (25)  1 (12) | 2 (20)  2 (20)  1 (10)  2 (20)  1 (10)  1 (10)  0 (0) |

IMD Decile: index of multiple deprivation decile, where 1 is most deprived and 10 is least deprived; SEN: on school educational needs register; SFA: small for age; LAC: ‘looked after child’, i.e. under care of local authority; prevLAC: previously looked after, i.e. adopted; FASD=Fetal alcohol Spectrum disorder; ASD: Autism spectrum disorder; ADHD: Attention deficit Hyperactivity disorder; DLD: Developmental Language disorder.

**Supplementary Table 1.2:** Descriptive data for Cases of FASD and Possible FASD from four schools excluded from the main study

|  | **FASD** | | **Possible FASD** | |
| --- | --- | --- | --- | --- |
|  | (FASD5) | (FASD6) | (POSS5) | (POSS6) |
| **Reason for invite** | SEN | LAC + SEN | SEN | PARENT OPT IN |
| **PAE reported** | Y | Y | Y | Y |
| **Domain with deficit** |  | | | |
| **Growth** | N | N | N | N |
| **Face** | Y | Y | N | N |
| **Neurological hard/soft** | Y | Y | Y | Y |
| **Communication** | Y | Y | Y | Y |
| **Cognition** | N | Y | Y | Y |
| **Executive function** | Y | Y | Y | N |
| **Memory** | N | Y | N | N |
| **Attention** | Y | Y | Y | Y |
| **Adaptive skills** | Y | Y | Y | Y |
| **Academic** | Y | Y | Y | N |

SEN: on school educational needs register; LAC: under the care of local authority. Y: meets criteria; N: does not meet criteria; PAE: prenatal alcohol exposure.
