## Supplementary file 2 for "Prevalence of Fetal Alcohol Spectrum Disorder (FASD) in Greater Manchester, UK: an active case ascertainment study"

**Supplementary Table 2.1:** Descriptive data for cases and possible cases of Fetal Alcohol Spectrum Disorders (FASD) in Schools 1-3 (main study)

|  | **FASD** | | | | **Possible FASD** | | | |
| --- | --- | --- | --- | --- | --- | --- | --- | --- |
|  | (FASD1) | (FASD2) | (FASD3) | (FASD4) | (POSS1) | (POSS2) | (POSS3) | (POSS4) |
| **Reason for invite** | SFA + SEN | LAC | Parent Opt-in | SFA + PrevLAC | SEN | SEN | SEN | SEN |
| **PAE reported** | N* | Y | Y | Y | Y | Y | Y | Y |
| **Domain with deficit** |  | | | | | | | |
| Growth | Y | N | N | Y | N | N | N | N |
| Face | Y | N | N | N | N | Y | N | N |
| Neurological hard/soft | N | Y | Y | Y | Y | Y | Y | Y |
| Communication | Y | Y | Y | Y | Y | Y | Y | Y |
| Cognition | Y | N | N | N | N | N | N | N |
| Executive function | Y | N | Y | Y | Y | Y | Y | N |
| Memory | Y | N | N | Y | N | Y | N | N |
| Attention | Y | Y | N | Y | N | Y | Y | N |
| Adaptive skills | Y | Y | Y | Y | Y | Y | N | Y |
| Academic | Y | N | N | N | Y | Y | Y | Y |

SFA small for age; SEN on school educational needs register; LAC under the care of local authority; PrevLAC adopted. Y meets criteria; N does not meet criteria. (* Child met criteria for FAS diagnosis, PAE disclosure not required)

POSS1: prenatal alcohol exposure as reported did not quite meet high risk PAE criteria but there were significant deficits in five Central Nervous System (CNS) domains and a normal microarray.

POSS2: missing microarray, therefore it was not possible to rule out genetic causes, however all other conditions were fully met including significant facial features.

POSS3: microarray was normal but all CNS deficits were borderline (only just met the threshold), and the prenatal alcohol exposure reported was vague (and deemed unreliable by the interviewer and case conference panel review).

POSS4: the threshold deficiency in three CNS sub domains was met and microarray was normal but facial features and Full Scale Intelligence Quota (FSIQ) were normal. The prenatal alcohol exposure reported was vague (and deemed unreliable by the interviewer and case conference panel review).
